# Effectiveness and Safety of Combining Tofacitinib with a Biologic in Patients with Refractory Inflammatory Bowel Diseases

**DOI:** 10.1101/2020.10.18.20214841

**Authors:** Quazim A. Alayo, Aava Khatiwada, Anish Patel, Maria Zulfiqar, Anas Gremida, Alexandra Gutierrez, Richard P. Rood, Matthew A. Ciorba, George Christophi, Parakkal Deepak

## Abstract

**Background:** One therapeutic option with limited data among patients with active moderate-to-severe ulcerative colitis (UC) and Crohn’s disease (CD) despite biologic monotherapy is using a combination of a biologic with Tofacitinib (TBT). Our aim was to examine the effectiveness and safety of TBT in this subset of patients.

**Methods:** Data of IBD patients at 2 referral centers on TBT were extracted. The primary outcome was clinical response (>50% reduction in symptoms) at week 8 and/or 16 determined by Physician Global Assessment. Secondary outcome was clinical remission (resolution of symptoms), corticosteroid-free clinical response and remission, normalization of CRP and endoscopic/radiographic response and remission. Adverse events (AEs) including any abnormal lipid profile or surgical complications were also assessed.

**Results:** Thirty-five patients (25UC, 10CD) were included. Biologics combined with tofacitinib were vedolizumab (68.6%), ustekinumab (17.1%), and infliximab (14.3%) and the median follow-up duration was 4 months. A majority (57.2%) had failed at least two biologics prior to starting TBT. At weeks 8 and/or 16, 37.1% achieved clinical response with 5.7% in clinical remission. Among the 23 patients with endoscopically/radiographically active disease at baseline, 56.5% had endoscopic/radiographic response and 34.8% achieved remission. Three AEs occurred in 2 (5.7%) patients, with an IR of 20.5 (15.0–47.2)/100PYF. No VTE and herpes zoster was reported.

**Conclusions:** TBT is effective at inducing endoscopic/radiographic response and a modest clinical response in UC and CD patients with active clinical symptoms despite prior biologic monotherapy. No new safety signals were detected beyond those reported with tofacitinib monotherapy.

## INTRODUCTION

Inflammatory bowel diseases (IBD), composed of Crohn’s disease (CD) and ulcerative colitis (UC), are a group of immune-mediated diseases with increasing prevalence and morbidity.^1^ Over the last two decades, the discovery and development of biologics and small molecules has expanded the therapeutic options for patients with moderate to severe IBD. Despite these novel therapeutics, there remain a substantial proportion of patients who either fail to respond, lose response or develop antibodies to these therapeutics.^2^ These patients often end up dependent on prolonged steroid use, having frequent disease flares and hospitalizations, undergoing multiple surgeries, and having a low quality of life.^3^ There is therefore a need for alternative therapeutic approach to managing these patients.

Based on the prior studies demonstarting that a combination of a biologic with immunomodulator is more effective than immunomodulator alone, there has been an increasing attempt to explore the therapeutic potential of combining mechanistically different biologics and/or small molecules in refractory IBD patiennts.^4^ However while the efficacy and safety of each individual biologics in IBD is well established, the efficacy and safety of a combinatorial therapy of two biologics or a small molecule and a biologic is yet to be shown in randomized control trials. Recently, there have been an increasing number of retrospective case series demonstrating that a dual biologic therapy (DBT) combining mechanistically different biologics may be a potential therapeutic option for refractory IBD patients,^5-8^ there is however paucity of data on combination of biologic with a small molecule.

One small molecule that has gained increased interest as a therapeutic option for severe inflammatory diseases including IBD is tofacitinib. Tofacitinib is an oral, small-molecule Janus Kinase (JAK) inhibitor, preferentially inhibits JAK1 and JAK3 which regulate signaling for multiple immune-relevant mediators implicated in the pathogenesis of inflammatory bowel diseases.^9, 10^ Tofacitinib has been approved for the treatment of moderate to severe UC based on safety and efficacy data from multiple clinical trials.^11, 12^ Although tofacitinib has not been approved for CD due to lack of demonstrable clinical efficacy in two phase II clinical trials,^13, 14^ a post-hoc analysis of these studies has shown some clinical benefit and biologic activity of tofacitinib in moderate to severe CD without new safety signal.^15^ Compared to tofacitinib, biologics such as the tumour necrosis factor-α (TNF) antagonists (infliximab, adalimumab and certolizumab pegol), antibodies inhibiting interleukins (IL)-12 and −23 (ustekinumab) and α4 β 7 integrins on gut-seeking leukocytes (vedolizumab) have mechanistically different targets in the IBD inflammatory pathway. Therefore combination of these biologics with tofacitinib may provide a synergistic effect on efficacy.

Here, we present the effectiveness and safety of combining tofacitinib, an oral, small-molecule Janus Kinase (JAK) inhibitor, with various biologics in patients with refractory IBD.

## METHODS

### Study design and settings

We conducted a retrospective review of all IBD patients with **T**ofacitinib added to other **B**iologic **T**herapy (TBT) at 2 IBD referal centers in the US (Washington University in St. Louis School of Medicine, St. Louis, MO, and Brooke Army Medical Center, Fort Sam Houston, TX).

### Participants

Patients were included if they were prescribed and used tofacitinib concomitantly with any other biologic for the treatment of UC or CD, and had at least one follow up visit. All patients had failed at least one biologic and were started on TBT because of active bowel-related clinical symptoms that had not responded to prior conventional therapy. Patients who were started on TBT primarily for an active extraintestinal manifestation were excluded. Data were collected retrospectively at each site between December 2018 and May 2020.

### Variables

Data collection was performed using a standardized data collection form on REDCap (Research Electronic Data Capture, version 7.3.5: REDCap Consortium, Vanderbilt University, Nashville, TN, U.S.A.), using pre-specified definitions and criteria for coding. Baseline demographic, clinical, endoscopic, radiographic and laboratory data were collected prior to initiation of TBT. Follow up assessments were done at weeks 8, 16, 26, 39 and 52 (+/- 4 weeks). Clinical response (assessed based on Physician Global Assessment (PGA)), corticosteroid or immunomodulator use, laboratory data, adverse events, IBD-related hospitalizations, herpes zoster reactivation, venous thrombo-embolism (VTE), major adverse coronary events (MACE) and IBD-related surgeries and complications were recorded at each timepoints. Disease activity was recorded from endoscopy (presence of mucosa ulcerations, Mayo endoscopic subscore and simple endoscopic score for CD [SES-CD]) and magnetic resonance enterography (MRE) scans (Global simplified magnetic resonance index of activity [MaRIAs]), performed within a year prior to starting combination therapy and at any time point during follow up.

### Outcomes

The primary effectiveness outcome was clinical response (>50% reduction in symptoms or remission) at week 8 and/or 16. The secondary effectiveness outcomes icluded corticosteroid-free clinical response, clinical remission (resolution of symptoms), corticosteroid-free clinical remission, endoscopic/radiographic response or remission, and normalization of CRP. Endoscopic/radiographic response or remission were determined using the aforementioned scores as previously described.^16-18^ Endoscopic/radiographic response was defined as a ≥1grade reduction in Mayo endoscopic subscore (UC), or > 50% reduction in SES-CD or ≥ 50% reduction in the Global MaRIAs score. We defined endoscopic/radiographic remission as follow up Mayo subscore of 0 or 1 (UC), Simplified SES-CD of 0-2 (CD) or Global MaRIAs score of < 6 (CD). Adverse events (AEs) were defined as serious if life-threatening or resulting in a hospitalization, disability or discontinuation of therapy. Abnormal lipid profile was defined as the presence of any one of the following: total cholesterol ≥200, low density lipoprotein (LDL)≥130, high density lipoprotein (HDL) <40 or Triglycerides ≥150.^19^ Surgical complications of abdominal surgery were graded using the Clavien-Dindo Classification and Comprehensive Complication Index.^20^

### Statistical analysis

We presented descriptive statistics as medians with interquartile range (IQR) or range for continuous variables and frequencies and percentages for categorical variables. Non-parametric continuous and categorical variables were compared using Wilcoxon rank-sum and Fisher’s exact tests respectively. Incidence rates (IRs) were calculated based on the unique number of patients with events per 100 patient-years of exposure. Exact Poisson CIs (adjusted for patient-years) are provided. All data were analyzed based on observed values, with no imputation for missing values. Statistical tests were two-sided and *p*-value of <0.05 was considered statistically significant. Analyses and graphically representations were done using Stata version 16.0 (StataCorp, College Station, TX) and GraphPad Prism version 8.3.0 (GraphPad Software; La Jolla, California, USA) respectively.

### Ethical Considerations

The study was approved by Institutional Review Boards (IRB) of both institutions and was conducted in accordance with the Declaration of Helsinki. This study is reported according to the Strengthening the Reporting of Observational Studies in Epidemiology (STROBE) statement for cohort studies.^21^

## RESULT

### Baseline characteristics

Thirty-five IBD patients (25UC, 10CD) were included in this study with a median follow-up of 4 months (IQR 2.6-5.9). The median age at initiation of TBT was 32 years (IQR 26 - 40). Majority were female (n=18, 51.4%), non-Hispanic white (23, 65.7%), and never smokers (26, 76.5%) and the median BMI of the cohort at baseline was 24.9 (22 – 29) kg/m^2^. Disease extent in the UC cohort was most frequently left-sided or distal (17/25, 69%), and disease location among the CD cohort was mostly ileocolonic (8/10, 80%) with 50% (5/10) having a non-stricturing/non-penetrating Montreal phenotype. More than half (20, 57.2%) had failed 2 or more biologics prior to starting TBT, and all patients received an induction dose of 10mg BID tofacitinib. Of the 30 patients who went on maintenance tofacitinib therapy, most (20, 66.7%) were on 5mg BID or 11mg XR daily dosing. The biologics combined with tofacitinib were vedolizumab (VDZ-TBT, n=24, 68.6%), infliximab (IFX-TBT, 6, 17.1%) and ustekinumab (UST-TBT, 5, 14.3%). A minority of patients (2, 5.7%) used concurrent immunomodulator at the time of initiation of TBT while 46.1% (16) were on concurrent corticosteroids (**Table 1)**

**Table 1:** Baseline Demographics and Clinical Characteristics.

| Characteristics |  | N=35 |
| --- | --- | --- |
| Age in years, median (IQR) |  | 32 (26 – 39) |
| Male, n (%) |  | 17 (48.6) |
| Race, n (%) | Non-Hispanic white | 23 (65.7) |
|  | African American/Black | 9 (25.7) |
|  | Hispanic | 1 (2.9) |
|  | Asians | 2 (5.7) |
| Body mass index kg/m<sup>2</sup>, median (IQR) |  | 24.9 (22 – 29) |
| Smoking Status, n (%)<br>N=34 | Never Smoker | 26 (76.5) |
|  | Past or Current Smoker | 8 (23.5) |
| Charlson comorbidity index, median (IQR) |  | 0 (0-0) |
| IBD type, n (%) | Ulcerative colitis | 25 (71.6) |
|  | Crohn's Disease | 10 (28.6) |
| UC extent, n (%)<br>N= 25 | E1: proctitis | 6 (23.0) |
|  | E2: left-sided or distal (till splenic Flexure) | 17 (69.0) |
|  | E3: extensive UC (proximal/splenic Flexure) | 1 (4.0) |
|  | Not reported | 1 (4.0) |
| CD Montreal location, n (%) N=10 | Ileal | 1 (10.0) |
|  | Colonic | 1 (10.0) |
|  | Ileocolonic | 8 (80.0) |
| CD Montreal Phenotype, n (%)<br>N=10 | Non-stricturing/ Non-penetrating | 5 (50.0) |
|  | Stricturing | - |
|  | Penetrating | 5 (50.0) |
| Upper GI disease in CD patients, n (%) N=10 |  | 1 (10.0) |
| Perianal disease in CD patients, n (%) N=10 |  | 6 (60.0) |
| Number of prior biologics, median (IQR) |  | 2 (1 - 3) |
| Number of prior biologics failed, n (%) | 1 | 15 (42.8) |
|  | 2 | 10 (28.6) |
|  | ≥3 | 10 (28.6) |
| Induction dosing, n (%) | 10 mg bid | 35 (100) |
| Maintenance dosing, n (%) N= 30 | 5mg bid or 11mg daily | 20 (66.7) |
|  | 10mg bid | 10 (33.3) |
| Biologic combined with Tofacitinib, n (%) | Infliximab | 6 (17.1) |
|  | Ustekinumab | 5 (14.3) |
|  | Vedolizumab | 24 (68.6) |
| Pre-TBT CRP (mg/dL), Median (IQR) |  | 1.35 (0.5-11.6) |
| Pre-TBT endoscopic/radiographic scores, median (IQR) | Mayo endoscopic sub-score (N=22) | 3 (2 – 3) |
|  | SES-CD (N=6) | 20.5 (9 – 24) |
|  | Global MaRIAs (N=4) | 2.5 (1.5 – 3) |
| Concurrent corticosteroid usage at combination start, n (%) |  | 16 (45.7) |
| Concurrent immunomodulator usage at combination start, n (%) |  | 2 (5.7) |
| Duration of IBD prior to starting tofacitinib in years, median (IQR) |  | 9 (3 – 16) |
| Follow up in months, median (IQR) |  | 4.0 (2.6 – 5.9) |
| Total exposure, Patient-years of follow up (PYF) |  | 14.9 |

Bid, twice daily; CD, Crohn's disease; GI, Gastrointestinal; IQR, Interquartile range; MaRIAs, simplified magnetic resonance index of activity; PYF, Patient-year of follow-up; SES-CD, Simple endoscopic score-Crohn's disease; TBT, Tofacitinib plus Biologic Therapy; UC, Ulcerative colitis

### Clinical Assessment

Physician global assessment data was available for all patients at 8/16 weeks from initiation of TBT with 13 patients (37.1%) achieving clinical response and 2 patients (5.7%) in clinical remission. At weeks 8/16, corticosteroid-free clinical response was achieved in 25.7% (9) and corticosteroid-free clinical remission was seen in 5.7% (2) of patients. Clinical response reported separately at weeks 8, 16 and 26 is shown in **Supplementary table S1**. At last documented assessment, clinical response was observed in 54.3% (n=19) while 45.7% (16) either had no response (14) or had lost response (1) **(Supplementary table S1)**. Clinical response at 8/16 weeks and at last documented follow up visits based on IBD type and combination biologic type is shown in **figures 1a and 1b**. Twenty percent (n=4) of patients on VDZ-TBT had clinical response at week 8/16 while the corresponding percentages in IFX-TBT and UST-TBT patients were 60.7 (n=3) and 50% (n=4) respectively **(Figure 1b)**. There was no significant difference in clinical response between patients who had previously failed 1 biologic and those who had failed 2 or more biologics prior to starting TBT (8/16 weeks, p=0.74; last documented follow up, p=0.31) **(Figure 1c)**. A univariate comparison of baseline characteristics of patients based on clinical response at 8/16 weeks was only significantly different for race (non-Hispanic whites vs. others, p=0.01) **(Table 2)**. TBT was discontinued in 20 patients (57.5%), most commonly due to no response (15, 75%). Other reasons TBT was discontinued were loss of response (1, 5%), de-escalation to monotherapy following clinical response on TBT (1, 5%), developing antibodies to the biologic (1, 5%), patient’s concern about susceptibility to COVID-19 with TBT (1, 5%), and loss to follow up ater 3 months of combinations therapy (1, 5%). The median persistence on tofacitinib was about 4.4 months (18.7 weeks) **(Figure 1d)**.

**Table 2:** Comparison of Baseline Characteristics based on Clinical Response at Week 8 or 16 on Tofacitinib plus Biologic therapy in IBD Patients.

|  |  | Clinical response at 8/16weeks |  | P values |
| --- | --- | --- | --- | --- |
|  |  | No (N=22) | Yes (N=13) |  |
| Age at tofacitinib induction, median (IQR) |  | 32.5 (26 – 42) | 31 (27 - 35) | 0.49 |
| Male, n (%) |  | 12 (54.5) | 5 (38.5) | 0.49 |
| Race, n (%) | Non-Hispanic white | 11 (50.0) | 12 (92.3) | **0.01** |
|  | Others | 11 (50.0) | 1 (7.7) |  |
| BMI, median (IQR) |  | 22.7 (20.9 – 30) | 25.3 (23.4 – 28.3) | 0.40 |
| IBD duration in years, mean (IQR) |  | 9.5 (3 - 19) | 3 (1 - 12) | 0.08 |
| Smoking Status<sup>†</sup>, n (%)<br>N=34 | Never Smoker | 17 (77.3) | 9 (75.0) | 0.88 |
|  | Current or past smoker | 5 (22.7) | 3 (25.0) |  |
| Concurrent steroid use at start of tofacitinib, n (%) |  | 12 (44.5) | 4 (30.7) | 0.29 |
| Concurrent immunomodulator use at start of tofacitinib, n (%) |  | 0 (0) | 2 (15.4) | 0.13 |
| Baseline hemoglobin (g/dL) median (IQR), N=33 |  | 12.1 (11.4 -13.5) | 11.4 (10.4-13.7) | 0.56 |
| Baseline albumin (g/dL), median (IQR), N=31 |  | 3.9 (3.5 – 4.2) | 4.0 (3.4 – 4.2) | 0.84 |
| Baseline CRP (mg/dL), median (IQR), n=26 |  | 0.6 (0.4 – 1.4) | 0.6 (0.2 – 3.8) | 0.67 |
| No of biologic class prior to tofacitinib, n (%) | 1 | 10 (45.4) | 5 (38.5) | 0.74 |
|  | ≥2 | 12 (54.5) | 8 (61.5) |  |

<sup>†</sup>Smoking status unknown in 1 patient
Abbreviations: bid, Two times a day; BMI, body mass index; CD; Crohn's disease; IBD-U, Inflammatory bowel disease unclassified; IQR, Interquartile range; PYF, Patient years follow up

**Figure 1:**
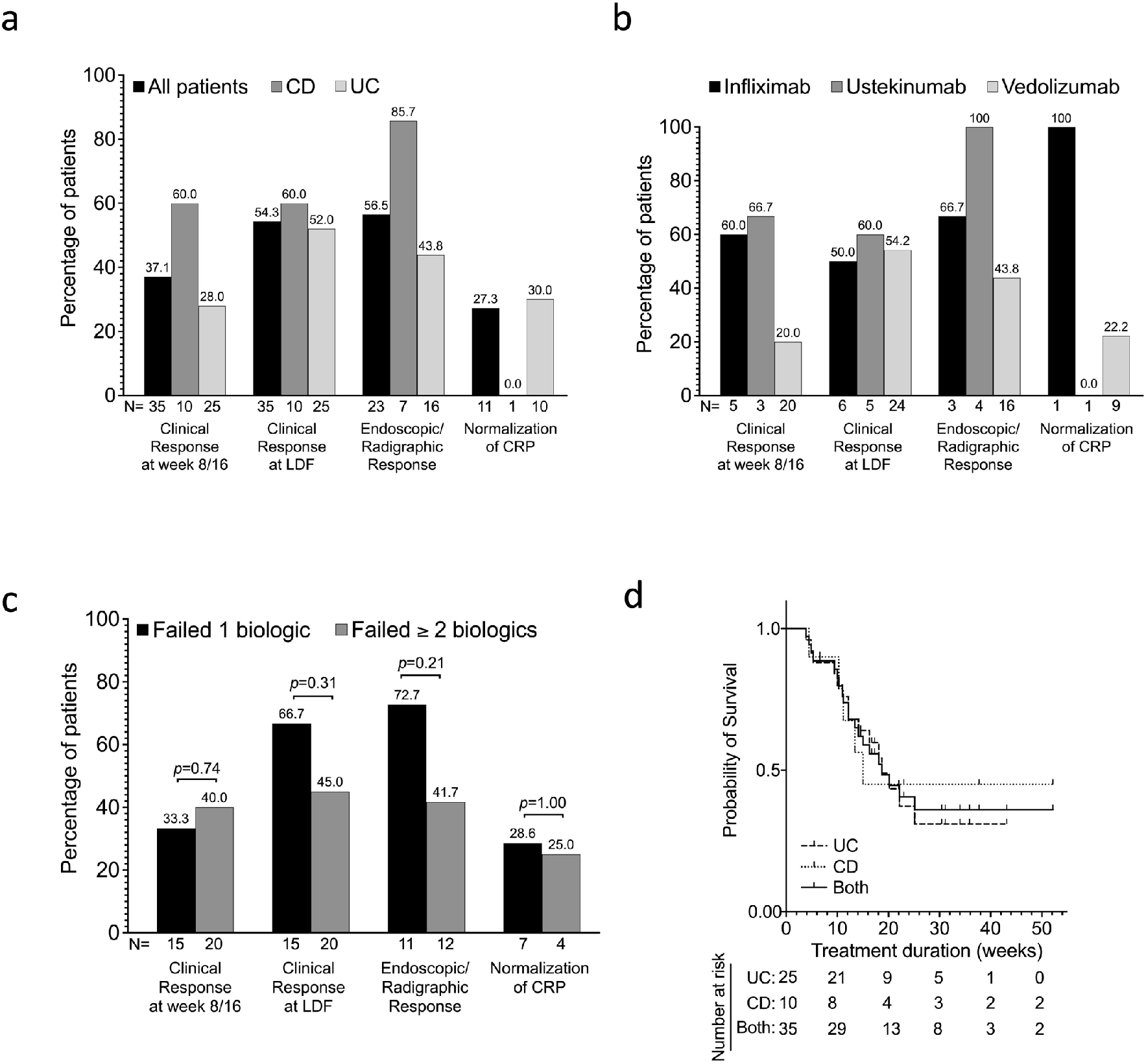
Effectiveness of tofacitinib plus Biologic Therapy (TBT) in moderate to severe IBD - clinical, endoscopic/radiographic and laboratory data. **A and B)** Bars showing the percentages of patients who had clinical response at week 8 and/or week 16, clinical response at last day of follow up (LDF), endoscopic/ radiographic response (among those with abnormal endoscopy/radiography at baseline) and normalization of CRP (among those with elevated CRP at baseline) while on TBT based on **A)** type of IBD (CD vs. UC) and **B)** type of combinatorial therapy (IFX-TBT vs. UST-TBT vs. VDZ-TBT). N represents the total number of patients in each subgroup. **C)** Association between number of prior failed biologics and the different outcomes in all patients shown in figure 1A. Bars represents proportion of patients in each outcome subgroup who failed one biologic and those who failed two or more biologics. Proportions were compared using Fisher exact test, and *P*-values are shown. N represents the total number of patients in each subgroup. **D)** Kaplan-Meier survivor curve of TBT persistence among UC (25), CD (10) and all patients (Both, 35) during the first year of treatment. Failure event was defined as withdrawal due to no response, loss of response or adverse events. All patients still on TBT as at week 52 of treatment were censored. The median times on tofacitinib for UC, CD and all patients (Both) were 18.7, 15.0 and 18.7 weeks respectively. CD, Crohn’s disease; CRP, C-reactive protein; IBD, Inflammatory bowel disease; IFX-TBT, Infliximab plus Tofacitinib; LDF, Last day of follow up, Ulcerative colitis, TBT, Tofacitinib plus Biologic Therapy, UST-TBT: Ustekinumab plus Tofacitinib; VDZ-TBT, Vedolizumab plus Tofacitinib

**Figure 2:**
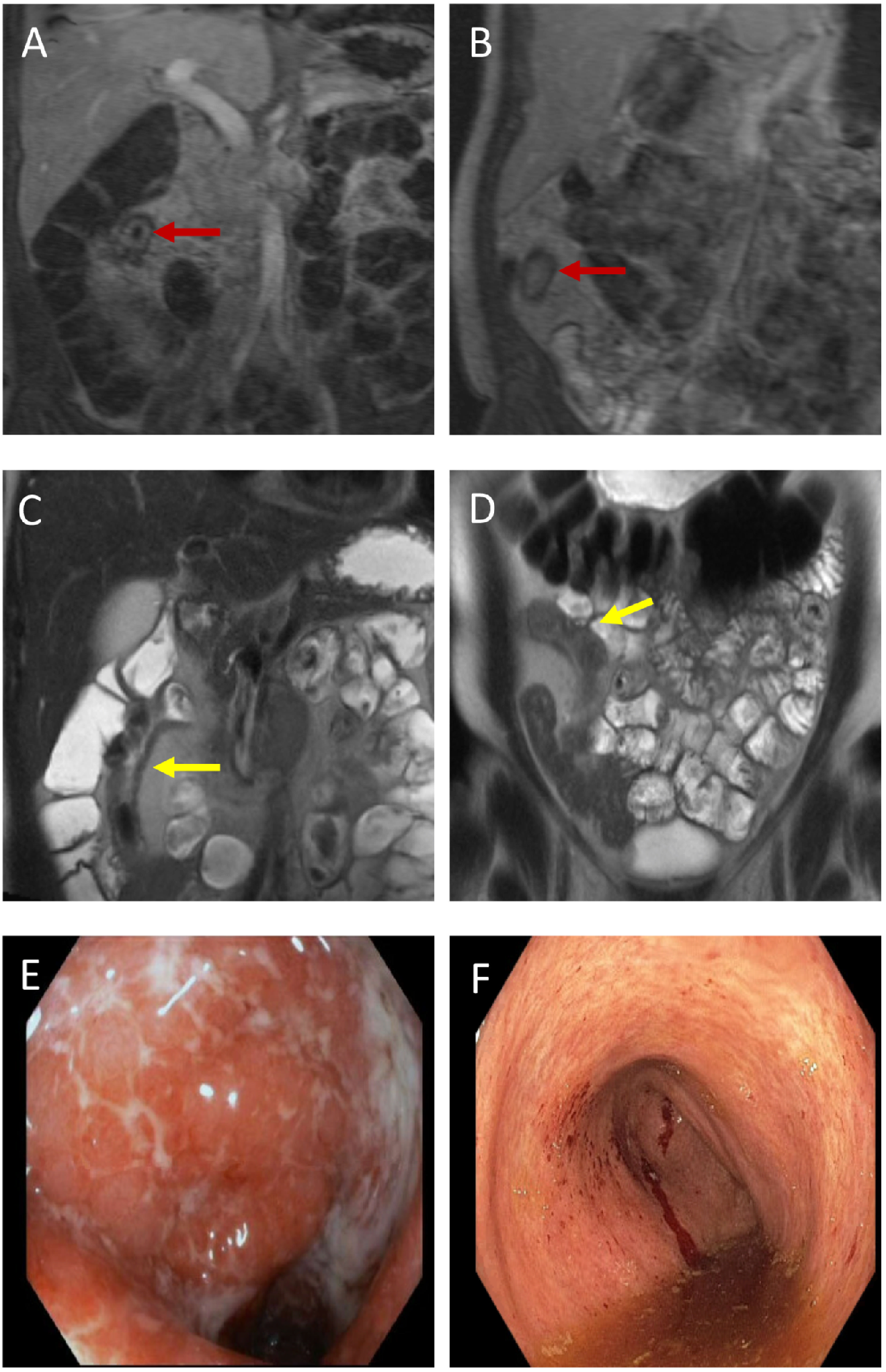
Representative magnetic resonance enterography (MRE) and endoscopic images of selected patients who had luminal response. Coronal MRE images in the portal venous phase prior to starting **(A)**, and while on tofacitinib plus biologic therapy (TBT) **(B)** demonstrate decreased interval enhancement of the inflamed terminal ileum in a patient with Crohn’s disease (red arrows). Coronal T2 HASTE MRE images of the same patient prior to starting **(C)**, and while on TBT **(D)** also show interval decreased wall thickness of the terminal ileum (yellow arrows). Endoscopic images of a different patient with ulcerative colitis prior to starting (**E**), and while on TBT (**F**) demonstrate interval decreased mucosal congestion and hyperemia representing treatment response. HASTE, Half-Fourier Acquisition Single-Shot Turbo-Spin-Echo; MRE, Magnetic Resonance Enterography

### Endoscopic/Radiographic Response

A total of 32 patients had a pre-TBT endoscopic/radiographic assessments (**Table 1**). Of these, 28 patients had pre- and on-TBT Mayo endoscopic subscore (21 UC patients), simple endoscopic score for CD (3 CD patients) and simplified magnetic resonance index of activity score (4 CD patients). Of the 23 patients who had endoscopically or radiographically active disease at baseline, 13 (56.5%) had an endoscopic/radiographic response while on TBT (**Figure 1a**), with 8 (34.8%) patients in endoscopic/radiographic remission. Endoscopic response by IBD type and combination biologic type is shown in **figures 1a and 1b**, limited to those who had endoscopically or radiographically active disease at baseline. Similar to clinical response, the number of prior failed biologics was not predictive of endoscopic/radiographic response. Not using concurrent systemic steroid at the start of TBT (61.5% vs. 10%, *p*=0.02) and a higher albumin level at baseline were predictive of an endoscopic/radiographic response (4.1g/dl vs. 3.5g/dl, *p*=0.001). Other baseline characteristics examined were not predictive of endoscopic/radiographic response **(Supplementary table S2). Figure 3** shows representative pre- and on-TBT MRE and endoscopic images of selected responders.

### Biochemical response

C-Reactive protein data at baseline and at week 8/16 were available for 20 patients in our cohort. Eleven (55%) of these patients had abnormal CRP at baseline. Normalization of CRP was achieved in 27.3% (n=8) of these patients at week 8/16. Biochemical response based on IBD type and combination biologic type is shown in **figures 1a and 1b**, limited to those with an elevated baseline CRP. Number of failed pre-TBT biologics did not predict biochemical response in this cohort (**Figure 1c**).

### Adverse Events

Three AEs occurred in 2 patients (5.7%) in our cohort with an incidence rate of 20.5 [95% Confidence interval (CI), 15.0-47.2] per 100 patient years follow-up (PYF) (**Table 3**). Both patients were male, had UC and were on 5 mg BID tofacitinib maintenance dosing. One patient on IFX-TBT developed Clostridium difficile infection and later developed candida esophagitis while on therapy. He was hospitalized for 8 days for the Clostridium difficile infection which was the only serious AE (IR 7.1 [95% CI, 5.0-10.4]/100PYF. The second patient was on VDZ-TBT and developed a rash. The median time to developing AEs was 32 days (Range 25-85). Combination therapy was not discontinued for any of these AEs and overall, no patient discontinued TBT due to adverse events. The patient who was hospitalized for Clostridium difficile infection later discontinued TBT 2 months after the infection due to loss of response. No DVT or herpes zoster reactivation or MACE was reported.

**Table 3:** Characteristics of Adverse Events.

|  |  |  |  |  |  |
| --- | --- | --- | --- | --- | --- |
| Total number of patients with an adverse event, n (%) |  |  |  | 2 (5.7) |  |
| Total number of patients with an serious adverse event, n (%) |  |  |  | 1 (5.7) |  |
| Incidence rate of all AEs, per 100 PYF [95%CI] |  |  |  | 20.5 (15.0 – 47.2) |  |
| Number of days to developing AEs, median [Range] |  |  |  | 32 [25– 85] |  |
| Adverse events | IBD type | Biologic | Tofacitinib maintenance dose | No of days on TBT | IR (/100PYF) |
| Clostridium difficile colitis*<sup>†</sup> | UC | Infliximab | 5mg BID | 25 | 6.8 |
| Candida esophagitis* | UC | Infliximab | 5mg BID | 85 | 6.8 |
| Rash | UC | Vedolizumab | 5mg BID | 32 | 6.8 |

\*Same patient developed Clostridium difficile colitis and candida esophagitis at different time points.
<sup>†</sup>Categorized as serious AE because this infection led to hospitalization while on TBT.
AE, Adverse event; IBD, Inflammatory bowel diseases; IQR, Interquartile range; PYF, Patient years follow-up.

### Lipid Profile

Paired lipid data at baseline and at 8/16 weeks was available for 21 patients. Of these, 9 (42.9%) had abnormal LP at baseline. One of 12 patients (7.14%) with normal lipid profile at baseline developed an abnormal profile at week 8/16 while 3 out of 9 patients (33.3%) with abnormal baseline lipids reverted to a normal profile at 8/16 weeks.

### IBD-Related Surgery and Hospitalization

Seven UC patients (20% of cohort) underwent total proctocolectomy plus IPAA and diverting ileostomy while on TBT (**Table 4**). All surgeries were due to failure of medical therapy and 2 (29%) were urgent or emergent. All surgical patients used tofacitinib within 2 weeks of surgery. At least one Clavien-Dindo grade complication was reported in 4 (33.3%) patients. Two patients had both a surgical site infection (SSI) and were on post-operative TPN. One patient had an SSI only and the last one required post-operative TPN. None had a Grade 3 complication. No patient had readmission or re-operation within 30 days of surgery. In addition to this, two other patients had a IBD-related hospitalization while on TBT. One was hospitalized for a Crohn’s disease flare and the other was hospitalized for clostridium difficile infection.

**Table 4:** IBD-related surgery and complications.

|  |  |
| --- | --- |
| Number of patients who underwent IBD-related surgery, n (% of cohort) | 7 (20.0) |
| **Timing of surgery:** |  |
| Urgent/emergent | 2 |
| Elective | 4 |
| Semi-elective | 1 |
| **Surgery indication:**<br>Refractory to medical therapy | 7 |
| **Surgery type:**<br>Total proctocolectomy, ileal pouch anal anastomosis, diverting ileostomy | 7 |
| **Surgical wound classification:**<br>Clean<br>Clean contaminated<br>Contaminated<br>Dirty infected | 7<br>-<br>-<br>- |
| Received tofacitinib within 2 weeks of surgery | 7 |
| Received tofacitinib within 4 weeks of surgery | 7 |
| Surgical Site Infection within 30 days of surgery | 3 |
| **Type of Surgical site infection**<br>Superficial incisional<br>Deep incisional | 3<br>- |
| Non-surgical/non-abdominal post-op infection | - |
| Post-op TPN | 3 |
| Experienced as least one Clavien-Dindo grade complication n (%) | 4 (50) |
| Experienced Clavien-Dindo Grade 3 or above complication | - |
| Steroid use prior to surgery | 4 |
| Re-operation within 30 days of surgery | - |
| Re-admission within 30 days of surgery | - |

Foot notes: IBD, Inflammatory bowel diseases; TPN, Total parenteral nutrition;

## DISCUSSION

Here, we report the effectiveness and safety of combining tofacitinib, a small molecule JAK inhibitor with biologics (TBT) in IBD patients who were refractory to prior biologic therapy. We showed that in a significantly refractory subset of IBD patients with a median disease duration of 9 years and more than half exposed to 2 or more prior biologics, 37.1% achieved clinical response on TBT at week 8/16, with 5.7% in corticosteroid-free clinical remission. We also showed among those with active disease on pre-TBT endoscopy/radiographic assessment, 56.5% achieved endoscopic/radiographic response and 34.8% were in endoscopic/radiographic remission while on TBT. Only 5.7% had an AE. No single case of HZ, DVT or MACE and no Clavien-Dindo complication of grade 3 or higher were reported among those who underwent surgery.

While there have been a number of recent reports using a combination of two biologics or a small molecule and a biologic in refractory IBD cases^5, 6, 8, 22-24^, most of these reports have been limited to DBT with only two case series and a case report reporting on TBT.^5, 25, 26^ Glassner et al. reported the overall effectiveness and safety of various combinatorial therapies in a cohort of 50 patients.^5^ Although, they reported the rate of AEs among the 20 patients on TBT in their cohort, the effectiveness in this subgroup was not separately reported. Clark-Snustad et al reported the rate of AEs in 18 patients on TBT for refractory CD without separately reporting the effectiveness in this cohort.^25^ Le Berre et al reported a case of a 67-year-old with left-sided UC who was successfully treated with a combination of tofacitinib and vedolizumab for refractory luminal and disabling rheumatologic disease.^26^ Our study therefore fills this gap in the knowledge of TBT by providing detailed data on its effectiveness and safety in moderate to severe IBD patients.

The rate of clinical response (28.0%) and clinical remission (4.0%) following induction in the 25 UC patients in our cohort were substantially lower than the corresponding rates reported in the prior induction clinal trials with 10mg BID of tofacitinib monotherapy in UC (average of OCTAVE induction 1 and 2: 57.5% clinical response,17.6% clinical remission). Similarly, the corresponding rates in the CD subgroup in our study (60% response and 10% remission) were lower than the prior clinical trials of tofacitinib monotherapy in CD (69.8% and 43.3% respectively). Notably, the overall clinical response (54.3%) and remission (22.9%) rate at last documented follow-up visits in our cohort were also low compared to what has been reported in DBT in IBD studies. Kwapisz et al. reported a clinical response of 73% in 15 refractory IBD patients on DBT^6^ while Yang et. al. reported 50% clinical response and 41% remission rates in their cohort of 22 CD patients on DBT.^8^ The baseline characteristics of these patients and the disparate methods of assessing clinical response and remission (PGA vs partial Mayo score/Crohn’s disease–patient-reported outcome-2 score [PRO2]) may account for these differences. More than 50% of our cohort had failed 2 or more biologic therapy while half of the patients in these clinical trials had only failed one prior biologic with the remaining half being biologic naïve.

Unlike the clinical response rate, endoscopic/radiographic response (56.5%) and remission (34.8%) induced by TBT in our patients were similar or slightly higher than what other retrospective DBT studies have reported (Kwapisz et al. - 44% in endoscopic/radiographic response; Yang et al. −43% in endoscopic response and 26% endoscopic remission).^6, 8^ This proportion is also similar to the sustained mucosal healing rate reported in the Octave Sustain trial of 5mg BID tofacitinib maintenance monotherapy in UC patients (27.8%). The congruence in endoscopic findings between our study and others may reflect the greater objectivity and lesser bias allowing the comparison of endoscopic/radiographic assessments between studies. Additionally, the poor correlation between clinical disease activity and endoscopic disease activity scores have been previously reported in the post-hoc analyses of the SONIC trial among patients with CD.^27^ Our study also showed that patients who were off systemic corticosteroids or those with higher albumin levels at baseline were more likely to have an endoscopic/radiographic response. These findings may suggest a lower disease activity among TBT responders compared to non-responders. Further prospective studies are needed to assess these and other baseline factors as predictive of a response to TBT.

A pooled analysis of seven DBT studies showed a higher proportion of AEs (38.9%) compared to our study (5.7%).^28^ Two other recent cases series of IBD patients on DBT by Yang et al. and Kwapicz et al. also reported higher rates of AEs (13% and 26.7% respectively) in their cohorts compared to our report, suggesting that TBT may be safer compared to DBT. Surprisingly, the proportion of patients with AE in our cohort was significantly lower than what was reported in prior clinical trials with tofacitinib monotherapy in UC (induction-54.9%, maintenance-79.6%) and CD (induction-54.5%, maintenance-79.6%).^11, 13^ Furthermore, the incidence rate of AE (20.7/100 PYF) reported here is lower than what we reported in our recently published real-world study of tofacitinib monotherapy in UC (IR: 27.2 (95% CI, 24.4–30.7) /100 PYF.^29^ Similarly, the proportion of patients with SAE in our TBT cohort is also lower compared to these previous tofacitinib monotherapy studies. This unexpected finding is likely due to differences in the definition and monitoring of AE between clinical trials and real-world data in our study as well as the relatively shorter duration of follow-up in the current study. However, head-to-head comparisons of TBT versus DBT in longer RCTs are necessary to fully assess the safety of TBT in comparison to DBT.

Tofacitinib has been associated with a dose-dependent risk of HZ in IBD patients and the FDA recently warned of increased risk of VTE and death when tofacitinib10mg BID is used in patients with rheumatoid arthritis (RA) and IBD.^30, 31^ However, none of our patients developed either of these complications while on TBT. We also did not find any case of MACE, new signal of lipid profile abnormalities and none of the 7 patients who underwent surgeries while on TBT experienced a severe Clavien-Dindo (grade 3 or higher) complication. Although, larger prospective studies with longer follow-up duration are needed to confirm these findings, our study adds to the growing body of evidence showing that DBT or combination of small molecule and biologics are well tolerated in IBD patients refractory to conventional biologic or small molecule monotherapy.

The strength of our study is that it is the largest cohort study to date from 2 large US IBD referral centers demonstrating the effectiveness and safety of TBT for luminal disease in a refractory IBD population. However, the retrospective study design, the assessment of clinical response using only the PGA, the possibility of missing AEs not adequately captured in the treating clinician documentation, lack of endoscopic follow-up in a small subset of patients, lack of a tofacitinib monotherapy control arm, and the short follow-up duration are notable limitations of our study. Despite these limitations, our results are similar to other reports on DBT or combination of small molecule and biologics in refractory IBD patients.

In conclusion, these results suggest that the combination of tofacitinib with a biologic agent induces a modest clinical response and significant endoscopic/radiographic response without any new safety signals in a subset of patients with IBD with active clinical symptoms despite biologic monotherapy.

## Data Availability

N/A

## Abbreviations

BID: Twice daily
CD: Crohn’s disease
COVID-19: Coronavirus disease 2019
CRP: C-Reactive protein
DBT: Dual biologic therapy
DVT: Deep venous thrombosis
HASTE: Half-Fourier Acquisition Single-Shot Turbo-Spin-Echo
HZV: Herpes Zozter Virus
IBD: Inflammatory bowel diseases
IFX-TBT: Infliximab plus Tofacitinib
IQR: Interquartile range
IR: Incidence rates
IRB: Institutional review board
JAK: Janus Kinase
MACE: Major adverse cardiovascular event
MRE: Magnetic resonance enterography
PGA: Physician Global Assessment
PYF: Patient-years of follow-up
SES-CD: Simple Endoscopic Score for Crohn’s Disease
sMARIA: Simplified Magnetic resonance index of activity
TBT: Tofacitinib plus Biologic Therapy
UC: Ulcerative colitis
UST-TBT: Ustekinumab plus Tofacitinib
VTE: Venous thromboembolism
VDZ-TBT: Vedolizumab plus Tofacitinib
XR: Extended release

## Notes

**Financial support:** Parakkal Deepak is supported by a Junior Faculty Development Award from the American College of Gastroenterology. Matthew A. Ciorba is supported by DK109384, a Crohn’s and Colitis Foundation Daniel H Present Senior Research Award (Ref. 370763) and philanthropic support from the Givin’ it all for Guts Foundation (https://givinitallforguts.org) and the Lawrence C. Pakula MD IBD Research Innovation and Education Fund. Anas Gremida is supported by the Lawrence C. Pakula, MD Advanced IBD Fellowship grant. Additional grant support for the REDCap database was provided by the Clinical and Translational Science Award (UL1 TR000448) and Siteman Cancer Center Support Grant (P30-CA091842).

### Competing Interest Statement

Quazim A. Alayo, Aava Khatiwada, Anish Patel, Maria Zulfiqar, Anas Gremida, George P. Christophi disclose no relevant conflicts of interest.
Parakkal Deepak: PD has served as a consultant or advisory board member for Janssen, Pfizer, Celgene, Arena, Prometheus; research grants from Takeda.
Alexandra Gutierrez: AG has served as a speaker for Janssen.
Richard Rood: RR is an advisor to Coloplast Corporation and has received research support from Gilead and Eli Lilly
Matthew A. Ciorba: MAC has done consultancy for Takeda, Pfizer and Prometheus labs and has been on the speaker bureau for AbbVie, Pfizer, Takeda, and UCB. Research support from Incyte. 

### Funding Statement

Parakkal Deepak is supported by a Junior Faculty Development Award from the American College of Gastroenterology. Matthew A. Ciorba is supported by DK109384, a Crohn's and Colitis Foundation Daniel H Present Senior Research Award (Ref. 370763) and philanthropic support from the Givin' it all for Guts Foundation (https://givinitallforguts.org) and the Lawrence C. Pakula MD IBD Research Innovation and Education Fund. Anas Gremida is supported by the Lawrence C. Pakula, MD Advanced IBD Fellowship grant. Additional grant support for the REDCap database was provided by the Clinical and Translational Science Award (UL1 TR000448) and Siteman Cancer Center Support Grant (P30-CA091842).

### Author Declarations

The study was approved by Institutional Review Boards (IRB) of both institutions and was conducted in accordance with the Declaration of Helsinki.

