## Supplementary Tables for "Effectiveness and Safety of Combining Tofacitinib with a Biologic in Patients with Refractory Inflammatory Bowel Diseases"

**Authors’ affiliations:**

1. Division of Gastroenterology and Inflammatory Bowel Diseases Center, Washington University in Saint Louis School of Medicine, St. Louis, MO, USA.
2. Department of Internal Medicine, St. Luke’s Hospital, Chesterfield, MO, USA.
3. Department of Medicine, Washington University in St. Louis School of Medicine, St. Louis, MO, USA.
4. Division of Gastroenterology, Brooke Army Medical Center, Fort Sam Houston, Texas, USA.
5. Department of Radiology, Mallinckrodt Institute of Radiology, Washington University in Saint Louis, St. Louis, MO, USA.
6. Department of Medicine, University of Central Florida, Orlando, FL, USA.

**Supplementary material**

**Supplementary table S1: Clinical response with TBT at various follow-up timepoints**

| Response assessments at different time points: | | | n (%) |
| --- | --- | --- | --- |
| Week 8, n (%),  N= 28 | Clinical response | Clinical remission | 2 (7.1) |
|  |  | 50% reduction in symptoms | 7 (25.0) |
|  | No response | | 19 (67.9) |
| Week 16, n (%),  N= 24 | Clinical response | Clinical remission | 1 (4.2) |
|  |  | 50% reduction in symptoms | 8 (33.3) |
|  | Loss of response | | 1 (4.2) |
|  | No response | | 14 (58.3) |
| Week 26, n (%),  N=10 | Clinical response | Clinical remission | 4 (40.3) |
|  |  | 50% reduction in symptoms | 5 (50.3) |
|  | No response | | 1 (8.3) |
| Response at last documented assessment, n (%), N= 35 | Clinical response | Clinical remission | 8 (22.9) |
|  |  | 50% reduction in symptoms | 11 (31.4) |
|  | Loss of response | | 1 (2.8) |
|  | No response | | 15 (42.9) |

Abbreviations: TBT, Tofacitinib plus Biologic Therapy

**Supplementary Table S2**: Comparison of Baseline Characteristics based on Endoscopic/Radiographic Response to TBT in IBD Patients with ulcerations at baseline

|  | | | | Endoscopic/Radiographic response | | P values |
| --- | --- | --- | --- | --- | --- | --- |
|  | | | | No (N=10) | Yes (N=13) |  |
| Age at tofacitinib induction, median (IQR) | | | | 30 (28 – 42) | 30 (24 - 33) | 0.32 |
| Male, n (%) | | | | 3 (30.0) | 6 (46.2) | 0.67 |
| Race, n (%) | Non-Hispanic white  Others | | | 6 (60.0)  4 (40.0) | 8 (61.5)  5 (38.5) | 1.00 |
| BMI, median (IQR) | | | | 22.5 (20.0 – 26) | 27 (22.7 – 28.8) | 0.28 |
| IBD duration in years, mean (IQR) | | | | 7.5 (3 - 12) | 7 (3 - 12) | 0.57 |
| Smoking Status, n (%) | Never Smoker  Current or past smoker | | | 7 (70.0)  3 (30.0) | 9 (69.2)  4 (30.8) | 1.00 |
| Maintenance dosing, n (%)  N=22 | 5mg bid/11mg daily  10mg bid | | | 8 (80.0)  2 (20.0) | 7 (59.3)  4 (41.7) | 0.27 |
| Concurrent steroid use at start of tofacitinib, n (%) | | No | | 1 (10.0) | 8 (61.5) | **0.02** |
|  |  | Yes | | 9 (90.0) | 5 (38.5) |  |
| Baseline hemoglobin (g/dL) median (IQR), N=21 | | | | 12.0 (11.4 -13.5) | 12.5 (11 - 13.8) | 0.82 |
| Baseline albumin (g/dL), median (IQR), N=22 | | | | 3.5 (3.3 – 3.7) | 4.1 (3.9 – 4.6) | **0.001** |
| Baseline CRP (mg/dL), median (IQR), n=19 | | | | 0.9 (0.5 – 1.8) | 0.5 (0.2 – 1.3) | 0.36 |
| No of biologic class prior to tofacitinib, n (%) | | | 1  ≥2 | 3 (30.0)  7 (70.0) | 5 (61.5)  8 (38.5) | 0.21 |

Abbreviations: bid, Two times a day; BMI, body mass index; CRP, C-reactive protein; IBD, Inflammatory bowel disease; IQR, Interquartile range; TBT, Tofacitinib plus Biologic Therapy
